# Lifestyle and Environmental Risk Factors Associated with Cancer: A Case-Control Study in Bangladesh

**DOI:** 10.1101/2025.07.09.25331165

**Authors:** Mohammad Lutfor Rahman, KM Tanvir, Farzana Rahman, Sreshtha Chowdhury, Shuvajit Saha, Md Abdullah Saeed Khan, Mohammad Azmain Iktidar, Mehedi Hasan, Samiha Nahar Tuli, Sharmin Akter, Tajrin Rahman, Mohammad Hayatun Nabi, Mohammad Delwer Hossain Hawlader

## Abstract

Cancer remains the second leading cause of death worldwide, with cases rising at an alarming rate. While the causes of cancer are complex and varied, certain risk factors—such as exposure to environmental pollutants and specific lifestyle choices—are modifiable and can be addressed. A case-control study, conducted from 25 August 2023 to 18 April 2024 in Bangladesh, explored the association between cancer risk and various lifestyle and environmental factors. The findings revealed that the use of wood or kerosene for cooking, consumption of improperly cooked fish and meat, exposure to cooking fumes, and frequent contact with inorganic dust, silica, and mosquito repellent significantly increase the risk of cancer. These results underscore the urgent need for public health initiatives focused on promoting healthier eating habits and reducing exposure to indoor air pollutants as key strategies for cancer prevention in Bangladesh.

## Introduction

Cancer is one of the potential causes of death worldwide, presenting a serious public health concern. The World Health Organization (WHO) reported that in 2018, Bangladesh experienced 150,781 new cases of cancer and 108,137 cancer-related deaths [1]. These alarming statistics highlight the pressing need for effective strategies to combat cancer, especially as projections suggest that both incidence and mortality rates are expected to increase in the coming decades. However, ongoing research underscores the potential for preventing a significant proportion of cancer cases through modifications in lifestyle and behavior-related risk factors [2].

Adopting a healthy lifestyle is crucial for the prevention and management of cancer, as well as other serious non-communicable diseases. Essential lifestyle changes include quitting smoking, which is a well-documented risk factor for various cancers, including lung and cervical cancers [3–5]. Maintaining a healthy weight is equally important, as obesity is linked to an increased risk of several cancers. Engaging in regular physical activity has been shown to significantly reduce the risk of breast cancer among women [6], while abstaining from excessive alcohol consumption can lower the risk of cancers such as those affecting the liver and esophagus. Additionally, a balanced diet rich in fruits, vegetables, and whole grains, while avoiding processed foods, contributes to cancer prevention [7].

Beyond lifestyle factors, reducing exposure to environmental pollutants is also critical. Both indoor and outdoor air pollution have been associated with an increased risk of cancer [8]. Pollutants such as particulate matter and industrial chemicals can contribute to cancer development. Moreover, excessive exposure to sunlight, radiation, and harmful chemicals found in some cosmetics has been linked to higher risks of skin cancer [9, 10].

Family history plays a significant role in cancer risk, with genetic predispositions heightening susceptibility to various types of cancer. Studies have demonstrated that individuals with a family history of cancer face elevated risks for prostate, gastric, colorectal, breast, and colon cancers [11–14]. Environmental factors such as air pollution, soil contamination, and temperature variations have also been positively correlated with increased cancer risk [15, 16]. Additionally, occupational exposures to hazardous substances have been shown to impact cancer incidence [17].

Despite these insights, there remains a research gap concerning the comprehensive effects of lifestyle factors and environmental exposures on cancer prevention and prognosis, particularly in Bangladesh. Specifically, a significant proportion of rural households in Bangladesh rely on traditional biomass fuels such as wood, cow dung, and kerosene for cooking. These fuels release harmful particulate matter and carcinogenic compounds into indoor environments, leading to prolonged exposure to toxic fumes.

Studies from other developing countries have shown a clear association between the use of biomass fuels and elevated risks of lung, gastrointestinal, and other cancers [18–21]. Given that around 60% (World Bank Report, 2024) of the Bangladeshi population lives in rural areas where this practice is widespread, it is crucial to investigate how these exposures influence cancer risk among this population.

Bengali cuisine often involves the consumption of fish and meat cooked at high temperatures or over an open flame. Improperly cooked foods, especially when charred or overcooked, can produce carcinogens such as heterocyclic amines and polycyclic aromatic hydrocarbons, which have been linked to cancer [22–24]. The cultural practice of cuisine distinguishes the Bangladeshi population from other ethnic groups, providing a critical area for investigation in cancer research.

Bangladesh’s rapid industrialization has led to increased exposure to various environmental pollutants such as inorganic dust, silica, and pesticides. Many rural Bangladeshis are exposed to these harmful environmental substances due to industrial and agricultural activities and poor occupational safety standards [25]. In addition, the use of mosquito repellents is widespread, especially during the monsoon season, and these repellents have been linked to increased cancer risk due to their chemical components [26]. The impact of these environmental exposures on cancer biology in the Bangladeshi population may differ from other regions due to both the intensity and duration of exposure.

Furthermore, there is a lack of extensive case-control studies that explore how adherence to specific lifestyle patterns—such as physical activity, dietary habits, and transportation modes—and exposure to environmental factors, including inorganic dust, cotton dust, wood dust, and silica, influence cancer risk. To address these research gaps, this study aims to update and synthesize existing knowledge on these associations. By focusing on epidemiological studies conducted in Bangladesh, we seek to provide a clearer understanding of how lifestyle and environmental factors contribute to cancer risk, ultimately informing more effective prevention and intervention strategies.

## Materials and methods

### Data

A case-control study was conducted to investigate the impact of dietary and environmental factors on cancer, spanning eight months from 25 August 2023 to 18 April 2024. The study involved patients diagnosed with six specific types of cancer: breast, hematological, oral, cervical, colorectal, and lung cancers. The patients were categorized as cases for having any one of these type of cancers. The controls were carefully selected to match the cases based on gender, age, and family background. Specifically, controls were chosen such that their year of birth was within 10 years of the cases to ensure age compatibility. Additionally, whenever possible, controls were selected from the same family as the cases to maintain genetic similarity.

A structured questionnaire was developed to capture relevant variables, including demographic factors (such as age, gender, educational qualification, and religion), lifestyle characteristics, environmental exposures, and dietary habits. This questionnaire was rigorously reviewed and refined by experts and was updated following a pilot survey conducted at National Insitute of Cancer Reserach and Hospital (NICRH)..

To determine the appropriate sample size for the study, we calculated an adjusted sample size of 350 cases based on established benchmarks. This sample size was deemed sufficient to address the primary research questions concerning the impact of dietary and environmental factors on cancer. Data collection was carried out across the country at 20 reputable cancer hospitals, which were randomly selected to provide a representative sample. Intern doctors, who were thoroughly trained prior to the study, were assigned to collect the data.

We successfully collected a total of 713 responses, with 361 from the case group. For each case, one control was intended to be matched; however, due to challenges in finding appropriate controls, nine cases did not have matched controls. Throughout the study, data collectors were closely supervised by two expert doctors to ensure the accuracy and reliability of the data. The collected data were then coded by a specialized data entry team.

### Variables

Dependent Variable: The dependent variable was binary, indicating whether a patient was diagnosed with one of six specific types of cancer: breast, hematological, oral, cervical, colorectal, or lung cancer. The presence of any type of cancer was considered a case and coded as 1, while the absence of cancer (controls) was coded as 0.

Independent Variables: There were two categories of independent variables: lifestyle and environmental factors. Lifestyle variables included type of transportation used, leisure time activity, type of cooking fuel, method of cooking meat, method of cooking fish, frequency of teeth cleaning, drinking water habit before breakfast, exposure to smoke during cooking, type of fish consumed, type of food consumed, amount of oil in food and frequency of cleaning or vacuuming at home. Environmental variables included exposure to engine exhaust, inorganic dust, cotton dust, silica, smog, pesticides, and mosquito repellent.

### Ethical Considerations

This study adhered to the ethical guidelines of Dhaka Ahsania Mission’s Institutional Review Board (IRB)/Ethical Review Committee (ERC) (Approval No.: 2023/DAM/IRB/ERC/2302). The ethical standards outlined in the 1964 Helsinki Declaration and its later revisions were followed where applicable. Written informed consent was obtained from all participants during face-to-face interviews, ensuring they understood their participation was voluntary and could be withdrawn at any time before signing.

### Statistical Analysis

The chi-square test was applied to assess the associations between categorical variables and cancer events. Multiple logistic regression was used to calculate adjusted odds ratios to evaluate the effects of dietary and environmental factors on cancer risk. All statistical tests were conducted with a significance level set at 5%. Statistical analysis was performed using Stata 17.

## Results

Table 1 presents the distribution of the selected characteristics for both cases and controls. In particular, educational levels differed significantly between the two groups: the majority of controls had higher levels of education, while the cases were more evenly distributed between various educational backgrounds. Furthermore, the study population was predominantly Muslim, with this characteristic being similarly distributed between both cases and controls. Other demographic and socioeconomic factors were also well distributed in cases and controls, indicating that the groups were comparable in these attributes. This distribution ensures that any observed differences in cancer risk factors are less likely to be attributable to discrepancies in these baseline characteristics.

**Table 1.**
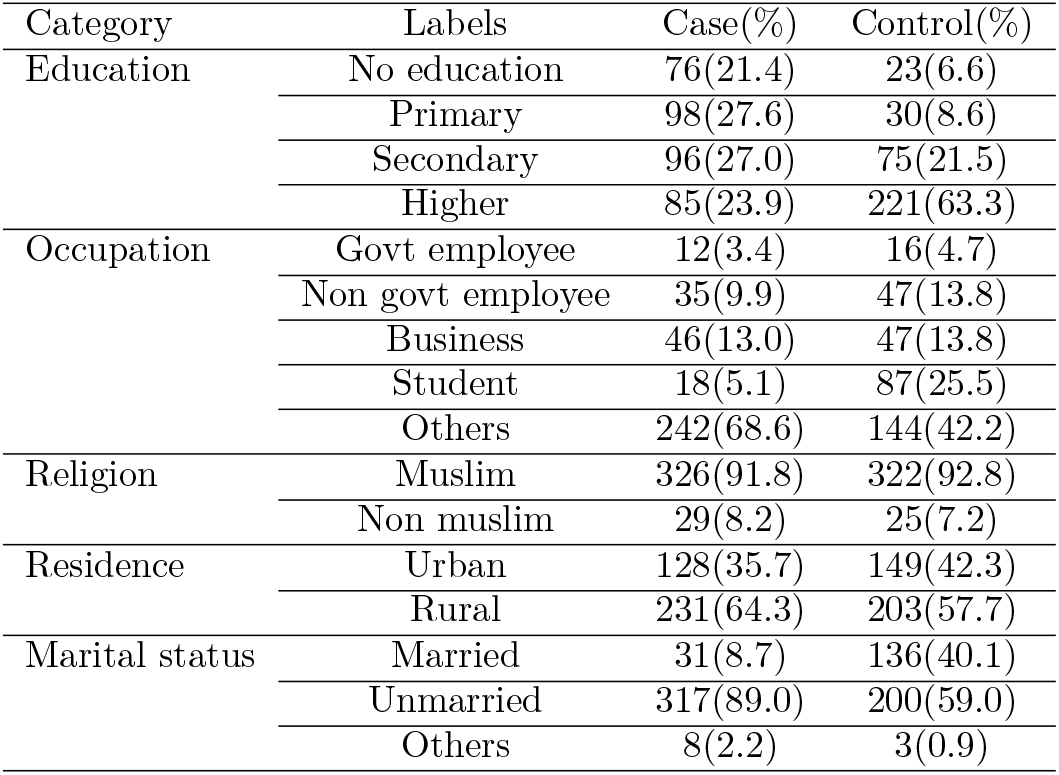
Selected characteristics of cases and controls (frequencies and percentages.

Table 2 details the distribution of individual lifestyle characteristics among cases and controls. Several key indicators related to lifestyle were significantly associated with cancer risk, including the type of fuel used for cooking, methods of cooking meat and fish, and exposure to smoke during cooking (p-values *<* 0.001).

**Table 2.**
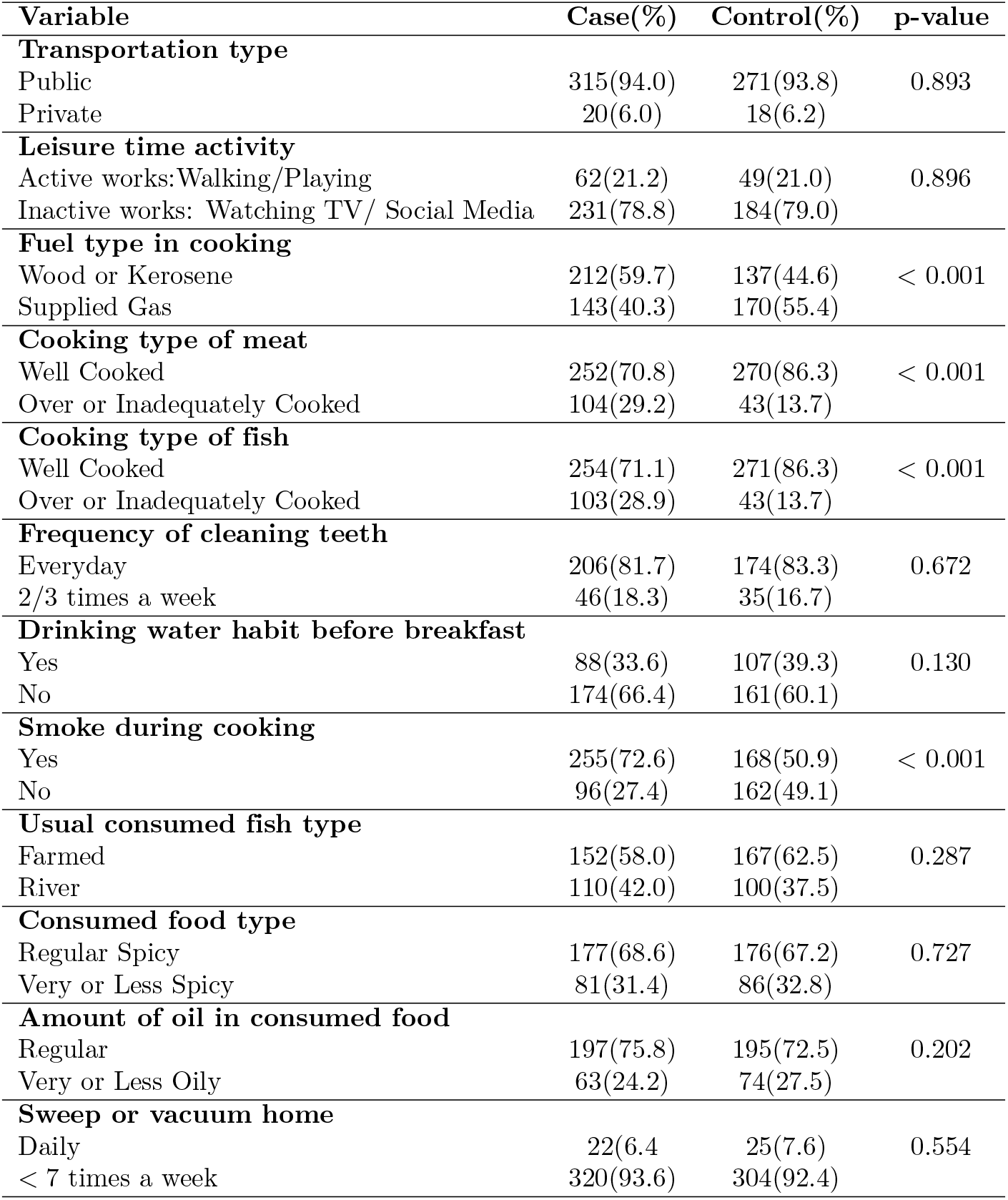
The lifestyle characteristics and their distribution among cases and controls.

| Variable | Case(%) | Control(%) | p-value |
| --- | --- | --- | --- |
| **Transportation type** |  |  |  |
| Public | 315(94.0) | 271(93.8) | 0.893 |
| Private | 20(6.0) | 18(6.2) |  |
| **Leisure time activity** |  |  |  |
| Active works:Walking/Playing | 62(21.2) | 49(21.0) | 0.896 |
| Inactive works: Watching TV/ Social Media | 231(78.8) | 184(79.0) |  |
| **Fuel type in cooking** |  |  |  |
| Wood or Kerosene | 212(59.7) | 137(44.6) | < 0.001 |
| Supplied Gas | 143(40.3) | 170(55.4) |  |
| **Cooking type of meat** |  |  |  |
| Well Cooked | 252(70.8) | 270(86.3) | < 0.001 |
| Over or Inadequately Cooked | 104(29.2) | 43(13.7) |  |
| **Cooking type of fish** |  |  |  |
| Well Cooked | 254(71.1) | 271(86.3) | < 0.001 |
| Over or Inadequately Cooked | 103(28.9) | 43(13.7) |  |
| **Frequency of cleaning teeth** |  |  |  |
| Everyday | 206(81.7) | 174(83.3) | 0.672 |
| 2/3 times a week | 46(18.3) | 35(16.7) |  |
| **Drinking water habit before breakfast** |  |  |  |
| Yes | 88(33.6) | 107(39.3) | 0.130 |
| No | 174(66.4) | 161(60.1) |  |
| **Smoke during cooking** |  |  |  |
| Yes | 255(72.6) | 168(50.9) | < 0.001 |
| No | 96(27.4) | 162(49.1) |  |
| **Usual consumed fish type** |  |  |  |
| Farmed | 152(58.0) | 167(62.5) | 0.287 |
| River | 110(42.0) | 100(37.5) |  |
| **Consumed food type** |  |  |  |
| Regular Spicy | 177(68.6) | 176(67.2) | 0.727 |
| Very or Less Spicy | 81(31.4) | 86(32.8) |  |
| **Amount of oil in consumed food** |  |  |  |
| Regular | 197(75.8) | 195(72.5) | 0.202 |
| Very or Less Oily | 63(24.2) | 74(27.5) |  |
| **Sweep or vacuum home** |  |  |  |
| Daily | 22(6.4) | 25(7.6) | 0.554 |
| < 7 times a week | 320(93.6) | 304(92.4) |  |

The findings reveal that a higher proportion of cases used wood or kerosene as cooking fuel (59.7%) compared to controls (44.6%). This suggests a notable association between the type of cooking fuel and cancer risk (p-values *<* 0.001). Additionally, 29.2% of cases consumed overcooked or inadequately cooked meat in contrast to 13.7% of controls (p-values *<* 0.001). Similarly, overcooked or raw fish consumption was reported by 28.9% of cases, whereas only 13.7% of controls reported this dietary habit (p-values *<* 0.001).

Exposure to smoke during cooking was also significantly associated with cancer risk.

A higher percentage of cases were exposed to cooking smoke (72.6%) compared to controls (60.9%), highlighting a strong association between smoke exposure and cancer incidence. Other lifestyle factors considered in the study did not show significant associations with cancer risk. This indicates that the factors highlighted such as cooking fuel type, meat and fish preparation methods, and smoke exposure are particularly relevant in understanding the lifestyle and environmental contributions to cancer risk.

Table 3 presents the associations between various lifestyle factors and cancer risk after adjusting for demographic variables, revealing several significant relations. The type of cooking fuel used was notably associated with cancer risk; specifically, individuals using wood or kerosene for cooking had 4.781 times odds of developing cancer compared to those using supplied gas (95% CI = 2.103–10.862), suggesting that the use of less clean cooking fuels may be linked with as increased cancer risk.

**Table 3.** Multivariate-adjusted odds ratios and confidence intervals of cancer prevalence associated to the lifestyle characteristics.

| Category | Labels | Adjusted OR | p-value | 95% CI |
| --- | --- | --- | --- | --- |
| **Transportation type** | Public<sup>reference</sup> |  |  |  |
|  | Private | 0.604 | 0.555 | (0.113, 3.219) |
| **Leisure time activity** | Active works<sup>reference</sup> |  |  |  |
|  | Inactive works | 2.683 | 0.267 | (0.469, 15.347) |
| **Fuel type in cooking** | Supplied Gas<sup>reference</sup> |  |  |  |
|  | Wood or Kerosene | 4.78 | < 0.001 | (10.862, 2.103) |
| **Cooking type of meat** | Well Cooked<sup>reference</sup> |  |  |  |
|  | Over or Inadequately Cooked | 1.640 | 0.562 | (0.308, 8.728) |
| **Cooking type of fish** | Well Cooked<sup>reference</sup> |  |  |  |
|  | Over or Inadequately Cooked | 1.213 | 0.828 | (0.212, 6.924) |
| **Frequency of cleaning teeth** | Everyday<sup>reference</sup> |  |  |  |
|  | 2/3 times a week | 6.045 | 0.001 | (2.010, 18.183) |
| **Drinking water habit before breakfast** | Yes<sup>reference</sup> |  |  |  |
|  | No | 1.242 | 0.615 | (0.534, 2.886) |
| **Smoke during cooking** | Yes<sup>reference</sup> |  |  |  |
|  | No | 0.332 | 0.019 | (0.132, 0.834) |
| **Usual consumed fish type** | Farmed<sup>reference</sup> |  |  |  |
|  | River | 0.620 | 0.335 | (0.235, 1.639) |
| **Consumed food type** | Regular Spicy<sup>reference</sup> |  |  |  |
|  | Very or Less Spicy | .936 | 0.879 | (0.400, 2.192) |
| **Amount of oil in consumed food** | Regular<sup>reference</sup> |  |  |  |
|  | Very or Less Oily | 0.328 | 0.015 | (0.133, 0.807) |
| **Sweep or vacuum home** | Daily<sup>reference</sup> |  |  |  |
|  | < 7 times a week | 1.697 | 6.589 | (0.437, 6.589) |

Additionally, oral hygiene practices were significantly related to cancer risk, with individuals who brushed their teeth 2–3 times a week facing six times higher odds of cancer compared to those who brushed daily (95% CI = 2.010–18.183), indicating the impact of consistent oral care on cancer risk. Dietary habits also played a role: those consuming very oily or less oily food had a 77.2% lower risk of cancer compared to those with regular oily food consumption (95% CI = 0.133–0.807), highlighting the potential benefits of reducing oil intake. Furthermore, exposure to smoke during cooking was significantly associated with cancer; individuals who were not exposed to cooking smoke had 66.8% lower odds of developing cancer compared to those who were exposed (95% CI = 0.132–0.834). These findings underscore the importance of addressing lifestyle factors such as cooking fuel type, oral hygiene, dietary oil content, and smoke exposure as part of strategies to reduce cancer risk.

Table 4 details the distribution of components of environmental exposures among cases and controls. Exposure to inorganic dust was significantly associated with cancer (p-value = 0.003) where 69.6% cases were cases and 58.0% were controls. Silica exposure also showed a strong association with cancer; 76.9% of cases were frequently exposed to silica, in contrast to 62.9% of controls, with the association being highly significant (p-value *<* 0.000).

**Table 4.** Environmental exposures and their distribution among cases and controls.

| Variable | Case(%) | Control(%) | p-value |
| --- | --- | --- | --- |
| **Engine exhaust** |  |  |  |
| Rarely | 155(49.4) | 173(54.7) | 0.264 |
| Frequently | 159(45.3) | 143(50.6) |  |
| **Inorganic dust** |  |  |  |
| Rarely | 94(30.4) | 133(42.0) | 0.003 |
| Frequently | 215(69.6) | 184(58.0) |  |
| **Cotton dust** |  |  |  |
| Rarely | 50(16.3) | 66(22.1) | 0.079 |
| Frequently | 256(83.7) | 232(77.9) |  |
| **Silica** |  |  |  |
| Rarely | 69(23.1) | 112(37.1) | <0.001 |
| Frequently | 230(76.9) | 190(62.9) |  |
| **Smog** |  |  |  |
| Rarely | 61(19.9) | 72(23.4) | 0.327 |
| Frequently | 246(80.1) | 236(76.6) |  |
| **Pesticides** |  |  |  |
| Rarely | 50(16.3) | 62(20.5) | 0.351 |
| Frequently | 257(83.7) | 240(79.5) |  |
| **Mosquito repellent** |  |  |  |
| Rarely | 146(43.6) | 206(65.4) | <0.001 |
| Frequently | 169(53.7) | 109(34.6) |  |

Additionally, the use of mosquito repellent was significantly linked to cancer risk, with 53.7% of cases frequently exposed to mosquito repellent compared to 34.6% of controls (p-value ¡ 0.000). Other environmental factors examined did not demonstrate significant associations with cancer at the 5% level of significance. These results highlight the substantial impact of inorganic dust, silica, and mosquito repellent exposure on cancer risk, emphasizing the need for targeted interventions in these areas.

Table 5 presents the adjusted odds ratios and confidence intervals for cancer prevalence associated with various environmental exposures taking into account the rarely exposed group as the reference. Our analysis indicates that exposure to mosquito repellent is significantly associated with an increased risk of cancer. Specifically, individuals frequently exposed to mosquito repellent have 79.8% higher odds of developing cancer compared to those who are rarely exposed to mosquito repellent (95% CI: 1.219-2.652). This significant association underscores the potential role of mosquito repellent exposure in elevating cancer risk.

**Table 5.** Multivariate-adjusted odds ratios and confidence intervals of cancer prevalence associated to the environmental factors.

| Category | Labels | Adjusted OR | p-value | 95% CI |
| --- | --- | --- | --- | --- |
| **Engine exhaust** | Rarely<sup>reference</sup> |  |  |  |
|  | Frequently | 0.908 | 0.663 | (0.588, 1.403) |
| **Inorganic dust** | Rarely<sup>reference</sup> |  |  |  |
|  | Frequently | 1.506 | 0.103 | (0.921, 2.463) |
| **Cotton dust** | Rarely<sup>reference</sup> |  |  |  |
|  | Frequently | 0.955 | 0.851 | (0.591, 1.543) |
| **Silica** | Rarely<sup>reference</sup> |  |  |  |
|  | Frequently | 1.321 | 0.219 | (0.848, 2.048 ) |
| **Smog** | Rarely<sup>reference</sup> |  |  |  |
|  | Frequently | 1.005 | 0.984 | (0.626, 1.613) |
| **Pesticides** | Rarely<sup>reference</sup> |  |  |  |
|  | Frequently | 1.030 | 0.908 | (0.621, 1.709) |
| **Mosquito repellent** | Rarely<sup>reference</sup> |  |  |  |
|  | Frequently | 1.798 | 0.003 | (1.219, 2.652) |

## Discussion

The current study investigated the association between lifestyle and environmental factors among cancer patients through a case-control study in Bangladesh. The primary objective was to identify potentially modifiable lifestyle risk factors that could help mitigate cancer prevalence. The study findings reveal significant associations between some lifestyle as well as environmental factors and cancer risk, providing valuable insights for public health interventions.

The present study found a significant association between the method of cooking meat and fish and cancer risk. Specifically, overcooking these foods was linked to an increased risk of cancer. This finding aligns with the European Food Safety Authority’s 2015 risk assessment, which indicated that overcooked food could elevate cancer risk across all age groups due to the formation of carcinogenic compounds such as heterocyclic amines and polycyclic aromatic hydrocarbons [27]. This evidence underscores the need for public awareness about safe cooking practices to reduce cancer risk.

In addition, we observed a significant relationship between cancer risk and the type of fuel used for cooking. Specifically, using wood and kerosene was associated with higher cancer risk. This is consistent with a study conducted in Iran, which reported that burning biomass or kerosene, especially in poorly ventilated areas, increased the risk of digestive cancers [20]. The use of such fuels contributes to indoor air pollution, which has been identified as a significant cancer risk factor. The current study also highlighted the adverse effects of cooking smoke on cancer risk. This finding is supported by research from Taiwan, which demonstrated that exposure to cooking fumes is associated with an increased risk of lung cancer [8]. Effective kitchen ventilation and smoke control are essential in reducing the potential health risks associated with cooking fumes.

Furthermore, this study identified a significant association between exposure to inorganic dust and silica and cancer risk. This is consistent with findings from Montreal, which linked exposure to these substances with an increased cancer risk [28]. The carcinogenic nature of inorganic dust and silica emphasizes the importance of implementing protective measures in environments where these exposures occur.

Finally, a significant association was found between cancer risk and the use of mosquito repellents. This finding is in agreement with research conducted in Taiwan, which reported a correlation between exposure to mosquito repellents and cancer risk [29]. The potential carcinogenic effects of some chemicals in mosquito repellents warrant cautious use and further investigation.

### Implications and Recommendations

The study findings suggest that various lifestyle and environmental factors influence significantly the cancer risk among Bangladeshis. These include the use of wood or kerosene as cooking fuel, exposure to cooking smoke, consumption of overcooked or raw food, and exposure to inorganic dust, silica, and mosquito repellents. Addressing these risk factors through targeted public health interventions could play a crucial role in reducing cancer incidence.

Public health initiatives should focus on promoting safer cooking practices, such as using cleaner fuels, improving kitchen ventilation, and reducing the consumption of overcooked foods. Additionally, minimizing exposure to hazardous substances like inorganic dust and mosquito repellents is essential. These measures could contribute to lowering cancer risk and improving overall public health [30].

Future research might be continued to explore the complex interactions between environmental, hormonal, and genetic factors in cancer development. Longitudinal studies and intervention trials would be valuable in understanding these relationships and developing effective cancer prevention strategies in Bangladesh.

## Data Availability

Data cannot be shared publicly at the moment because of confidentiality as we wish to continue further research in other aspects based on the current data. However, data will be available from the corresponding author via email subject to the approval of stakeholders.

## Acknowledgment

This study was supported by the Centennial Research Grant, established in celebration of the 100th anniversary of the University of Dhaka. We also extend our gratitude to dedicated public health researchers from the Department of Public Health of North South University for their invaluable contributions to questionnaire development, data collection, and project implementation.

## Author Contributions

**Conceptualization:** Mohammad Lutfor Rahman, KM Tanvir. **Data curation**: KM Tanvir, Mohammad Lutfor Rahman, Farzana Rahman. **Formal analysis:** KM Tanvir, Mohammad Lutfor Rahman. **Methodology:** Mohammad Lutfor Rahman, KM Tanvir, Project administration: Mohammad Lutfor Rahman. **Resources:** Farzana Rahman, Sreshtha Chowdhury, Shuvajit Saha, Mohammad Azmain Iktidar, Mehedi Hasan, Samiha Nahar Tuli, Tajrin Rahman. **Software:** KM Tanvir, Mohammad Lutfor Rahman. **Supervision:** Mohammad Lutfor Rahman, Mohammad Hayatun Nabi, Mohammad Delwer Hossain Hawlader. **Validation:** Mohammad Lutfor Rahman, Shuvajit Saha **Visualization**: Writing – original draft: KM Tanvir, Mohammad Lutfor Rahman **Writing – review & editing**: KM Tanvir, Mohammad Lutfor Rahman, Farzana Rahman, Shuvajit Saha, Md Abdullah Saeed Khan, Mohammad Hayatun Nabi, Mohammad Delwer Hossain Hawlader.

## Supporting information

**S1 Fig. Percentage of case and control according to lifestyle components**

**S2 Fig. Percentage of case and control frequently exposed to environmental components**

## Notes

### Competing Interest Statement

The authors have declared no competing interest.

### Funding Statement

Yes

### Author Declarations

Dhaka Ahsania Mission (DAM) is a leading non-governmental development organization established in 1958 by Khan Bahadur Ahsanullah, a distinguished educationist, social reformer, and Sufi spiritual leader. Guided by its founding motto—"Divine and Humanitarian Service"—DAM has been dedicated to the holistic development of society, encompassing both social and spiritual dimensions. As one of DAM’s three core sectors, the Health Sector plays a pivotal role in advancing community well-being through multifaceted programs. Its initiatives reflect a deep commitment to fostering positive change, ensuring equitable access to healthcare, and promoting a harmonious life for all. Through collaborative efforts with government agencies, national and international NGOs, donors, voluntary organizations, and compassionate individuals, the Health Sector continues to make significant strides in public health. To uphold ethical standards in research, DAM has established an Institutional Review Board (IRB) responsible for evaluating and approving studies involving human participants in Bangladesh. The current study has received ethical clearance from DAM’s IRB under the reference number: 2023/DAM/IRB/ERC/2302). With a legacy of dedicated service, DAM’s Health Sector remains steadfast in its mission to create a healthier, more equitable society—celebrating every milestone achieved through collective effort and unwavering determination.

